# MorphoStain: Physics-Guided Latent Diffusion and Morphological Distillation for Multiplex Stain Decomposition in Alzheimer’s Neuropathology

**DOI:** 10.64898/2026.11.15.26368147

**Authors:** Mohammadreza Saraei, Maryam Zand, Saba Rahmani

## Abstract

Multiplex immunohistochemistry (mIHC) enables simultaneous biomarker profiling, yet unmixing colocalized targets with identical spectra, such as dual-DAB marking Tau and Amyloid-*β* in Alzheimer’s pathology, remains mathematically ill-posed. We propose MorphoStain, an unsupervised, physics-guided framework for morphological stain decomposition with epistemic uncertainty quantification. MorphoStain models stain separation via a conditional Latent Diffusion Teacher operating on a Beer– Lambert optical-density manifold, guided by pathology foundation models. To enable real-time clinical throughput, the teacher is distilled into a FastStudentU-Net (0.12M parameters, *>* 1,400 *×* compression), while a Symmetric Morphological Routing Module dynamically disentangles identical chromogen overlap. Evaluated on 9,908 held-out patches from the SEA-AD cohort, MorphoStain achieves 40.56 dB PSNR and 0.9961 SSIM at 140.1 FPS, showing statistically significant gains over InverseAE (Cohen’s d = 3.52) and NMF (Cohen’s d = 7.02), with p *<* 0.001 for both comparisons.

## I. Introduction

**M**ULTIPLEX immunohistochemistry (mIHC) visualizes multiple molecular biomarkers within a single tissue section using distinct chromogens, enabling spatially resolved profiling with standard brightfield RGB scanners [1]–[4]. In Alzheimer’s disease (AD), co-staining amyloid-*β* (A*β*) and tau captures interacting pathologies, making accurate computational stain unmixing essential for downstream quantification. However, because plaques and neuritic structures are fine, weakly stained, and spatially adjacent, standard RGB sensors face a fundamental physical limit: under the Beer–Lambert model, optical density (OD) is a linear combination of stain absorption vectors [5], making the inversion of *m >* 2 overlapping stains ill-conditioned or strictly underdetermined.

Beyond this loss of identifiability, severe spectral crosstalk amplifies ambiguity. Precipitates like 3,3’-diaminobenzidine (DAB, brown) vary strongly in intensity and approach black at saturation, competing with the hematoxylin (blue) counterstain. Consequently, pixel-wise color matching [5]–[7] often misassigns signals or fragments fine structures. Resolving overlapping chromogens requires spatial morphology and learned priors (Fig. 1). While supervised deep models [8], [9] learn spatial context, they require costly annotations. Unsupervised methods like InverseAE [10]–[12] avoid annotations but remain deterministic, yielding a single point estimate without quantifying decomposition ambiguity across weakly stained plaques. Conversely, diffusion models [13]–[15] can model multi-modal posteriors and provide uncertainty estimates, but deploying them across gigapixel whole-slide images (WSIs) incurs prohibitive latency, and uncalibrated stochastic sampling may violate physical image-formation constraints.

**Fig 1:**
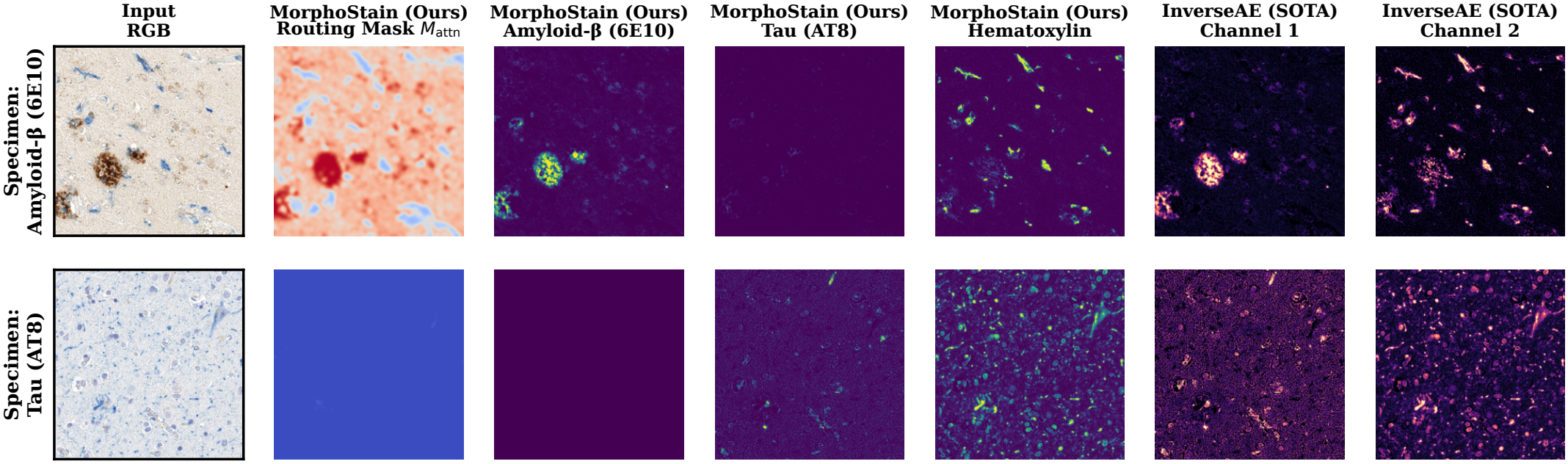
Qualitative comparison of unsupervised stain decomposition and biomarker disentanglement. Representative multiplex IHC neuropathology patches showing input RGB, MorphoStain symmetric routing maps (*M*_attn_), unmixed Amyloid-*β*, Tau (AT8), and Hematoxylin against the SOTA baseline.

To bridge this gap, we introduce MorphoStain, a physicsguided latent diffusion teacher paired with a compact distilled student for uncertainty-aware A*β*/tau stain unmixing. Our contributions are fourfold:

### Physics-Guided Latent Diffusion Teacher

We combine a calibrated Beer–Lambert projection (*S*_AD_ ∈ ℝ^3×2^) with a latent diffusion model conditioned on the Phikon pathology foundation model [16], using Monte Carlo sampling to estimate calibrated epistemic uncertainty (*u*_var_).

### Lightweight Student

Through knowledge distillation with dilated residual blocks (*d*∈ {1, 2, 4, 8} [17]–[19]), we achieve a *>* 1,400× compression (169.67M→ 0.12M parameters) for real-time WSI inference.

### Symmetric Morphological Routing (*M*_attn_)

A dedicated attention module that disentangles A*β* and tau morphologies, suppresses bleed-through, and preserves fine neuritic structures.

### Comprehensive Benchmarking

Extensive evaluation across 9,908 test patches, including zero-shot out-of-distribution (OOD) generalization and scanner illumination shifts [20], demonstrating promising reconstruction fidelity [21]–[23], morphological preservation, and uncertainty calibration.

## II. Related Work

### A. Classical Color Deconvolution

Classical unmixing operates in OD space via the Beer– Lambert law. Early methods rely on fixed OD vectors [5], singular value decomposition (SVD)-based plane estimation [6], or unsupervised matrix factorization [7], [24]–[26]. Because three RGB channels cannot uniquely resolve high-plex mixtures pixel-wise, sparse regression [27], [28] and groupsparsity priors [29] introduce regularization. However, lacking spatial context, pixel-wise methods frequently produce morphologically implausible decompositions under heavy spectral overlap, motivating our integration of physical forward models with deep structural priors.

### B. Deep Stain Decomposition

Deep networks leverage spatial context to overcome pixel-wise ambiguity [9], though weakly supervised methods still rely on manual point annotations [8]. The state-of-the-art (SOTA) unsupervised baseline, InverseAE [10], [11], trains a conditional autoencoder with an inverse-image reconstruction loss. Despite strong performance on general mIHC, InverseAE struggles with dark DAB saturation, assumes a single dominant stain per pixel, and smooths out high-frequency micro-textures. While generative adversarial network (GAN)-based unmixing [12] improves texture preservation, both approaches are strictly deterministic. In contrast, we model the full posterior distribution over physically valid decompositions specifically tailored to co-localized AD pathologies.

### C. Diffusion Models and Knowledge Distillation

Diffusion models [13], [15] and Latent Diffusion Models (LDMs) [14] excel at capturing complex multi-modal distributions, with recent adaptations like PathLDM [30] targeting histopathology synthesis. However, unconditional synthesis does not guarantee physical consistency with an observed RGB image, nor does raw sample variance guarantee calibrated uncertainty. We constrain latent diffusion with an explicit Beer–Lambert decoder and condition it on self-supervised pathology features from Phikon [16]. Finally, to overcome the high latency of iterative sampling on gigapixel WSIs, we distill our physics-informed teacher into a lightweight student for clinical-grade throughput.

## III. Methodology

Fig. 2 illustrates the proposed MorphoStain architecture. MorphoStain introduces a physics-guided teacher–student framework that couples classical Beer–Lambert OD principles with foundation-model-conditioned latent diffusion for morphological stain decomposition and epistemic uncertainty quantification in mIHC. To enable efficient inference, teacher-derived concentration distributions and epistemic uncertainty maps are distilled into a compact multi-branch student network (see Supplementary Algorithm S1 for the complete two-stage pseudocode).

**Fig 2:**
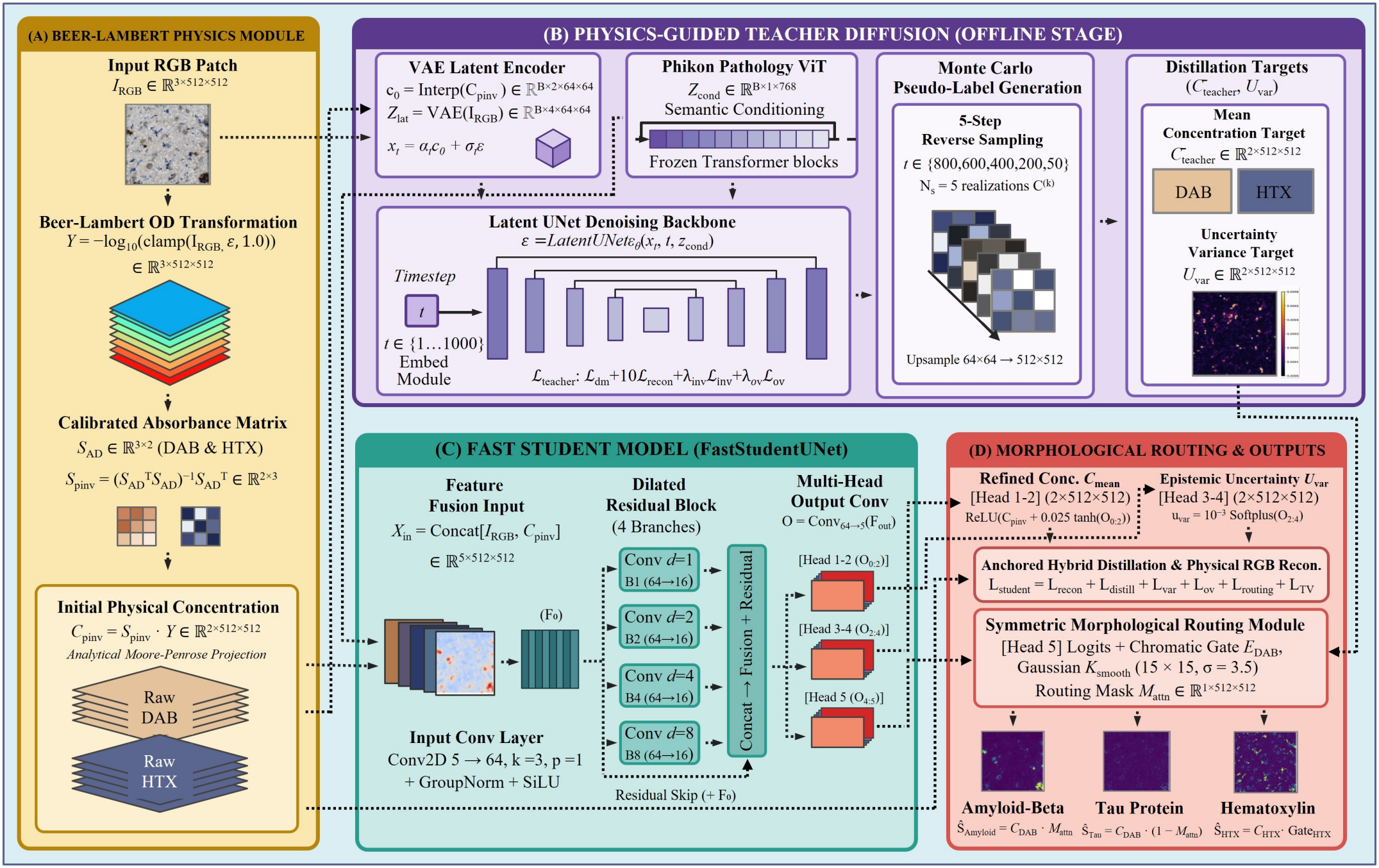
Architectural overview of MorphoStain. (A) Beer–Lambert Physics Module: Converts input patches *I*_RGB_ ∈ ℝ^3×512×512^ to optical density (OD) space *Y* = −log_10_(clamp(*I*_RGB_, *ϵ*, 1.0)) to derive initial physical concentrations *C*_pinv_ = *S*_pinv_ ·*Y* ∈ℝ^2×512×512^ via analytical Moore–Penrose projection using calibrated absorbance matrix *S*_AD_. (B) Physics-Guided Teacher Diffusion (Offline Stage): Operates on downsampled concentration priors *c*_0_ = Interp(*C*_pinv_) ∈ ℝ^*B*×2×64×64^ alongside variational autoencoder (VAE) spatial latents *Z*_lat_ ∈ ℝ^*B*×4×64×64^ conditioned on frozen Phikon pathology vision transformer (ViT) embeddings *Z*_cond_ ∈ ℝ^*B*×1×768^. Supervised via ℒ _teacher_, a five-step reverse sampling process (*N*_*s*_ = 5 realizations) followed by 64 × 64 →512 ×512 upsampling synthesizes pseudo-label ensembles, yielding mean concentration targets 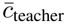 and epistemic uncertainty variance *U*_var_. (C) Fast Student Model: Ingests concatenated RGB-physics features *X*_in_ = Concat[*I*_RGB_, *C*_pinv_] ∈ ℝ^5×512×512^ through an initial convolution (*F*_0_) and four parallel dilated residual branches (*B*_1_, *B*_2_, *B*_4_, *B*_8_ with *d* 1, 2, 4, 8) to predict multi-head outputs *O* ∈ ℝ^5×512×512^. (D) Morphological Routing and Outputs: Aligns refined concentrations *C*_mean_ (Head 1–2) and epistemic uncertainty *U*_var_ (Head 3–4) via anchored hybrid distillation and physical RGB reconstruction (ℒ_student_), while a symmetric morphological routing module (Head 5 logits gated by chromatic envelope *E*_DAB_ and Gaussian smoothing *K*_smooth_, 15 ×15, *σ* = 3.5) generates routing mask *M*_attn_ to disentangle shared chromogens into Amyloid-*β* (*Ŝ*_Amyloid_), Tau (*Ŝ*_Tau_), and Hematoxylin (*Ŝ*_HTX_).

### A. Spectral Beer–Lambert Physics

Given an input RGB patch *I*_RGB_ ∈ ℝ^*B*×3×*H*×*W*^ (*H* = *W* = 512), the Beer–Lambert law [5] transforms non-linear RGB intensities into linear OD space via

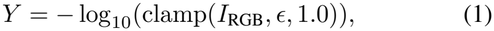

with *ϵ* = 10^−5^. In OD space, flattened observations *Y*_flat_ ∈ ℝ^3×*BHW*^ are modeled as a linear mixture *Y*_flat_ = *S*_AD_*C*_flat_, where *C*_flat_ ∈ ℝ^*M* ×*BHW*^ denotes local stain concentrations and *S*_AD_ ∈ ℝ^3×*M*^ is the column-wise *ℓ*_2_-normalized spectral absorbance matrix. For dual-target mIHC (*M* = 2: DAB brown for pathological protein aggregates and Hematoxylin blue for cell nuclei), we calibrate *S*_AD_ = Normalize(*S*_init_, *p* = 2, dim = 0) with

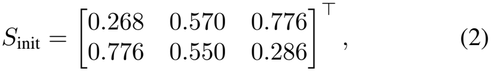

where columns represent DAB and Hematoxylin absorbance vectors, respectively. While *M <* 3 is nominally overdetermined, unmixing co-localized biomarkers under background noise and spectral bleed-through becomes ill-conditioned. We obtain an initial physical projection via the Moore–Penrose pseudoinverse 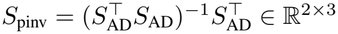, yielding

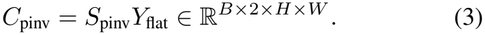

Conversely, any concentration map *C* is mapped back to normalized RGB space via exponential transmission:

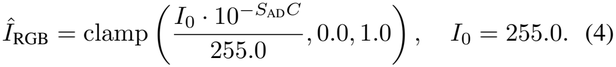

### B. Physics-Guided Teacher and Monte Carlo Pseudo-Labeling

#### 1) Latent Conditioning and Forward Process

To capture spatial histological priors without manual annotations, our teacher model (ℳ_1_) operates on a downsampled 64 × 64 concentration manifold *c*_0_ = Interpolate(*C*_pinv_, 64 × 64) along-side a pre-trained variational autoencoder (VAE) latent code *z*_lat_ ∈ ℝ^*B*×4×64×64^. To inject domain-specific pathological semantics, we extract conditioning embeddings from the Phikon foundation model [16]:

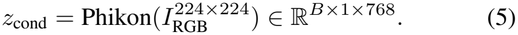

During forward diffusion across timesteps *t* ∼ Uniform(0, 1000), Gaussian noise *ϵ* ∼ N (0, *I*) is injected via

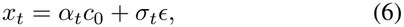

with *α*_*t*_ = 1 − 0.25(*t/*1000) and *σ*_*t*_ = 0.25(*t/*1000).

#### 2) Teacher Optimization

The denoising backbone predicts 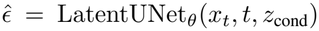. To prevent degenerate solutions in underdetermined color spaces, we optimize

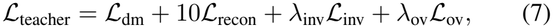

combining standard score matching 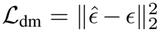 and RGB fidelity *ℒ* _recon_ = ∥ *Î*_RGB_ −*I*_RGB_ ∥ _1_ with two physical regularizers (*λ*_inv_ = 0.5, *λ*_ov_ = 0.10, *ϵ*_inv_ = 10^−5^):

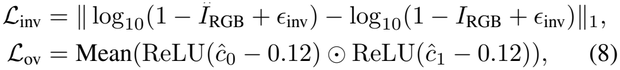

where *ℒ* _inv_ enforces absorption bounds in complementary transmission space and *ℒ* _ov_ penalizes implausible marker colocalization.

#### 3) Monte Carlo Pseudo-Labeling

To avoid costly iterative sampling at test time, ℳ_1_ generates offline pseudo-labels and epistemic uncertainty maps. For each training patch, we draw *N*_samples_ = 5 perturbed initializations 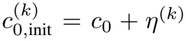 with *η*^(*k*)^ ∼ *N* (0, 0.20^2^*I*) and execute 5-step reverse denoising over *t* ∈ {800, 600, 400, 200, 50} via

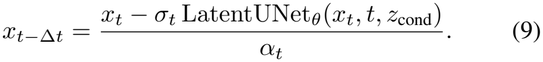

Upsampling the denoised latents to 512 × 512 yields full-resolution realizations *C*^(*k*)^, from which we compute pixel-wise target means and epistemic variances (∈ ℝ^*B*×2×512×512^):

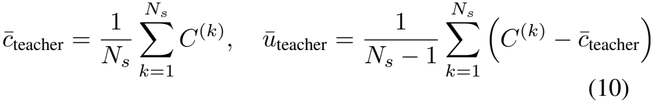

where *N*_*s*_ ≡ *N*_samples_.

### c. Distilled Student and Morphological Routing

#### 1) FastStudentUNet Architecture

To eliminate VAE and vision transformer (ViT) overhead during WSI inference, FastStudentUNet compresses the pipeline by *>* 1,400× (169.67M → 0.12M parameters). It takes a 5-channel physics-fused input *X*_in_ = Concat[*I*_RGB_, *C*_pinv_] ∈ ℝ^*B*×5×512×512^ and extracts initial features *F*_0_ = *ϕ*(Conv_5→64_(*X*_in_)), where *ϕ*(·) ≡SiLU(GN_8_(·)) denotes Group Normalization [31] and SiLU activation. To capture multi-scale context without grid artifacts [17]–[19], *F*_0_ passes through four parallel dilated 3×3 convolutions *B*_*d*_ = Conv_64→16,dil=*d*_(*F*_0_) for *d*∈{1, 2, 4, 8} , which are concatenated and fused residually:

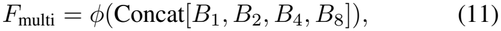

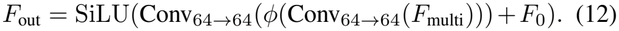

A final 3× 3 head predicts *O* = Conv_64→5_(*F*_out_) ∈ ℝ^*B*×5×512×512^, parameterized into refined concentrations, epistemic uncertainties, and smoothed routing logits:

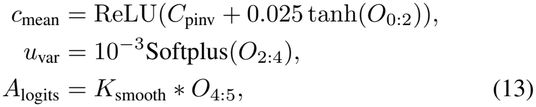

where *K*_smooth_ is a reflection-padded 15 × 15 Gaussian kernel (*σ*_gauss_ = 3.5).

#### 2) Symmetric Morphological Routing

Because A*β* plaques and Tau tangles share the DAB chromogen [1], we disentangle their co-localized signals using morphological attention gated by chromatic selectivity. We first isolate true DAB absorption from gray neuropil scattering via Δ_DAB_ = ReLU(*Y*_*B*_ − *Y*_*R*_) + 0.5 ReLU(*Y*_*G*_ − *Y*_*R*_) and compute a smooth chromatic envelope:

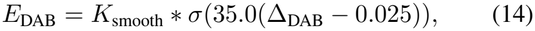

where *σ*(·) is the sigmoid function. The routing attention mask *M*_attn_ = *K*_smooth_ * (*σ*(*A*_logits_) ⊙*E*_DAB_) then partitions total concentrations *c*_mean_ = [*C*_DAB,total_, *C*_HTX_] into three disentangled channels:

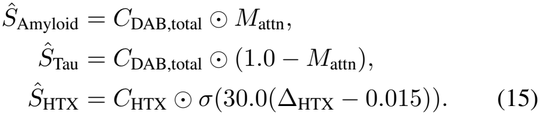

### D. Multi-Objective Student Distillation Loss

We train FastStudentUNet end-to-end by minimizing *ℒ*_student_ = *ℒ*_recon_ + *ℒ*_distill_ + *ℒ*_var_ + *ℒ*_ov_ + *ℒ*_routing_ + *ℒ*_TV_. Individual losses govern RGB fidelity, teacher distillation, error-aligned uncertainty calibration (using mean squared error map MSE_map_ = Mean_ch_((*Î*_RGB_ − *I*_RGB_)^2^) [21]), overlap suppression, and smooth morphological routing:

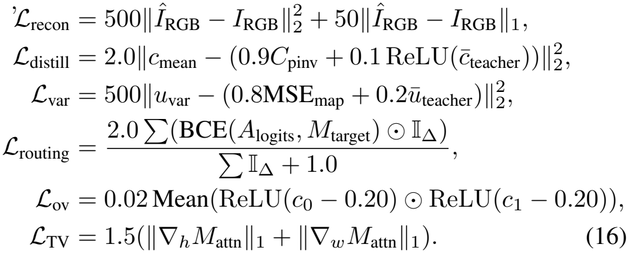

Here, *M*_target_ ∈{0, 1} is the slide-level biomarker label (0 for A*β*, 1 for Tau), and the indicator I_Δ_ = I(Δ_DAB_ *>* 0.04) restricts routing supervision strictly to active chromophore regions.

## IV. Experimental Setup

### A. Dataset and Preprocessing

We evaluate MorphoStain on 10 middle temporal gyrus (MTG) WSIs from 5 donors in the SEA-AD cohort [1], featuring paired tissue sections stained for Tau (AT8) and Amyloid-*β* (6E10) (Supplementary Tables S1 and S2). Slides were digitized using an Aperio scanner at 20× magnification (0.5016 *μ*m/pixel). Discarding non-overlapping 512 × 512 tiles with *<* 30% tissue coverage or optical defocusing yielded an initial 49,576 candidate patches. Following a secondary quality-control screening to remove sparse tissue or background artifacts, 37,538 high-quality patches remained. To prevent anatomical data leakage, these were partitioned into strictly isolated donor-level splits: H20.33.001/002/005 for training (*N*_train_ = 24,510), H19.33.004 for validation (*N*_val_ = 3,120), and H20.33.004 for testing (*N*_test_ = 9,908).

### B. Baselines and Implementation Details

We benchmark MorphoStain against seven representative unmixing methods spanning analytical, matrix factorization, and deep learning paradigms: classical color deconvolution (Ruifrok and Johnston [5]), SVD-based plane estimation (Macenko *et al*. [6]), pixel-wise *L*_1_-regularized regression (Las-soReg [27]), OD NMF [25], *L*_1_-regularized Sparse NMF [24], structure-preserving dictionary learning (Vahadane *et al*. [7]), and the SOTA unsupervised autoencoder InverseAE [11].

The Phikon-conditioned [16] teacher (Latent UNet) and student (FastStudentUNet) are both trained for 10 epochs using AdamW (*η* = 2 ×10^−4^ for teacher and 3 ×10^−4^ for student, with cosine annealing) using a batch size of *B* = 32. Training is conducted using mixed precision on a single NVIDIA RTX 6000 Ada (48GB) GPU. Grid-search sensitivity analyses for *λ*_ov_ and *K*_smooth_ are detailed in Supplementary Section S3 (Supplementary Fig. S1 and Supplementary Table S3). We assess RGB reconstruction fidelity via peak signal-to-noise ratio (PSNR), mean squared error (MSE), root mean squared error (RMSE) [21], and structural similarity index (SSIM) [22]; spectral alignment via Cosine Similarity [32]; perceptual texture preservation via LPIPS [23]; and computational efficiency via single-batch inference throughput (frames per second, FPS, at 512 × 512, *B* = 1).

## V. Benchmark Results and Analysis

### A. Comparative Quantitative Analysis

As detailed in Table I, MorphoStain sets a new standard for physical unmixing. Across the complete *N* = 9,908 held-out test cohort, it achieves a mean reconstruction PSNR of 40.56 dB and an SSIM of 0.9961 (see Supplementary Figs. S3 and S4 for qualitative unmixing across pathological ratios and the complete empirical PSNR density distribution). With these metrics, MorphoStain outperforms classical matrix decomposition baselines [5]–[7], [25] by margins of +5.17 dB to +19.16 dB (and by +22.19 dB over pixel-wise *L*_1_ regression [27]). Furthermore, it significantly surpasses the deep learning SOTA, InverseAE [11] (40.56 dB vs. 38.84 dB, a +1.72 dB gain).

Unlike InverseAE, which lacks morphological routing capabilities and consequently suffers catastrophic crosstalk bleed when resolving overlapping identical brown DAB chromogens into constituent Tau and Amyloid-*β* channels, MorphoStain successfully isolates these structures. Finally, MorphoStain achieves superior perceptual feature consistency (LPIPS = 0.0141) while providing real-time throughput at 140.1 FPS, capable of unmixing and stitching complete gigapixel WSIs (*>* 50,000 × 40,000 pixels) in under 12 s (Supplementary Fig. S6).

Crucially, paired patch-level hypothesis testing confirms that all performance gains over baselines are statistically significant (*p <* 10^−16^) with exceptionally large effect sizes (*e*.*g*., Cohen’s *d* = 3.52 vs. InverseAE, *d* = 7.02 vs. NMF). Patch-level variance reveals high consistency with a median PSNR of 40.52 dB and an interquartile range (IQR) of 1.42 dB (see Supplementary Tables S4 and S5).

### B. Ablation Analysis and Structural Distillation

We dissect architectural components by evaluating four teacher configurations (M1–M4) against the distilled student model on a standardized calibration subset (Table II; see Supplementary Fig. S2 for training convergence trajectories).

1. *Impact of Foundation Conditioning:* Removing Phikon ViT semantics [16] (M2) causes severe collapse (ΔPSNR = − 10.72 dB), establishing that global pathological embeddings are critical for unmixing guidance and convergence stability relative to the full teacher model (M1).
2. *Impact of Physics Constraints:* Omitting Beer–Lambert optical losses (M3) drops PSNR to 24.19 dB, confirming that unconstrained diffusion drifts into ill-posed local minima and fails to preserve realistic spectral unmixing. Similarly, excluding overlap penalties (M4) leads to severe chromophore bleed (PSNR = 27.21 dB).
3. *Student Distillation Dynamics:* FastStudentUNet delivers 41.66 dB PSNR and 174.6 FPS (5.73 ms latency) while compressing model complexity by *>* 1,400× (169.67M → 0.12M parameters), demonstrating that continuous convolution smoothing bypasses the sampling variance present in iterative reverse diffusion.

### C. Zero-Shot Out-of-Domain Generalization

Generalization was evaluated zero-shot across *N* = 100 unseen brain patches from an external validation cohort [1] (Table III), sampled from WSIs of donor case H14.09.010 spanning distinct neuropathological targets: Amyloid-*β* and phosphorylated Tau. To evaluate model stability and ensure strict reproducibility, all zero-shot evaluations were deterministically benchmarked across three independent random seeds (42, 123, 2024), aggregating a total of 300 evaluation runs.

As illustrated in Fig. 3, MorphoStain demonstrates strong domain transferability across diverse neuropathological morphologies (diffuse plaques and neurofibrillary tangles), maintaining 36.82 dB PSNR and 0.9808 SSIM (RMSE = 0.0148).

**Fig 3:**
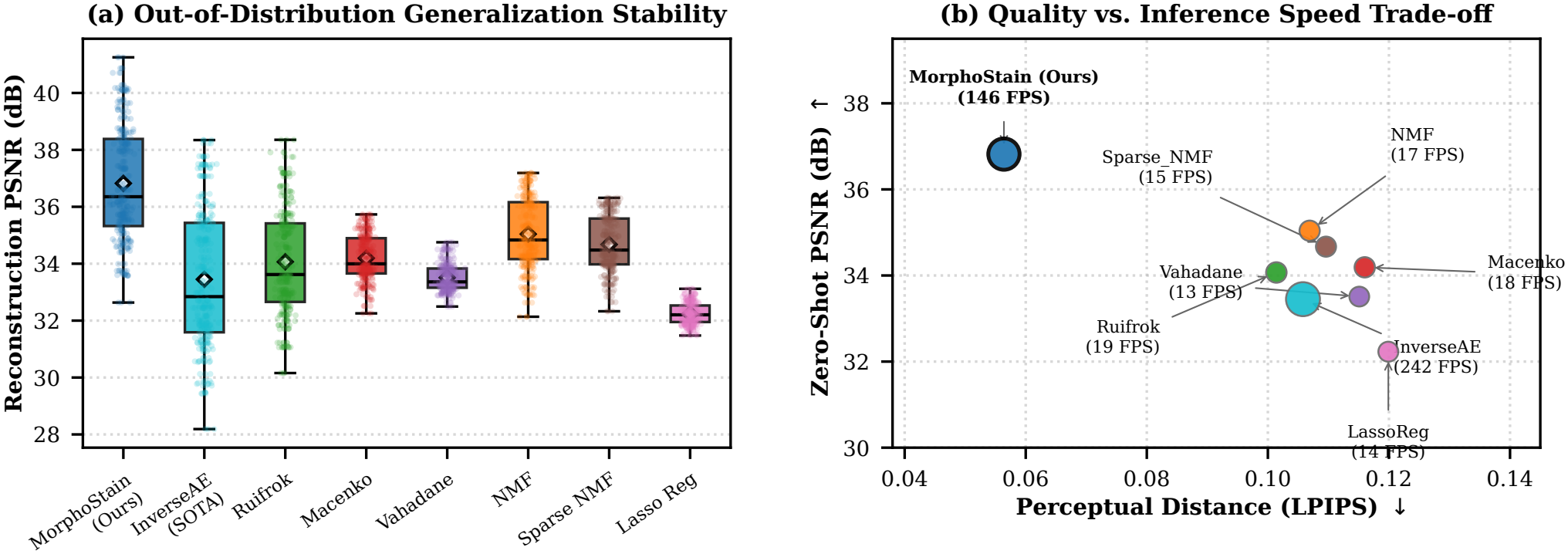
Zero-Shot Out-of-Distribution Validation. (a) Boxplot of PSNR distribution across 8 models. (b) Pareto chart evaluating quality versus computational efficiency (PSNR vs. LPIPS), where bubble size corresponds to inference speed (FPS).

It decisively surpasses both classical unmixing algorithms (+1.78 dB over NMF [25]) and the deep learning SOTA InverseAE (+3.37 dB gain), establishing an optimal Pareto frontier for visual quality and computational efficiency.

### D. Robustness to Illumination Drift and Sensor Noise

### 1) Illumination Drift

As reported in Table IV, under artificial illumination drifts up to +30% (Shift = 1.30), Mor-phoStain exhibits minimal degradation, retaining 35.37 dB PSNR (0.9776 SSIM) compared to InverseAE’s rapid collapse to 23.73 dB PSNR and 0.9325 SSIM (+11.64 dB margin; see Supplementary Fig. S8 for visual comparisons across progressive shift factors).

#### 2) Sensor Noise Tolerance

We evaluate resilience against additive Gaussian sensor noise across standard deviation scales *σ* ∈ [0.0, 0.10] (Table V and Fig. 4). Under nominal conditions (*σ* = 0.00), MorphoStain achieves a +1.30 dB advantage over InverseAE [11]. While both models exhibit comparable trajectories across moderate noise regimes (*σ* ∈ [0.01, 0.05]), MorphoStain maintains graceful degradation and mitigates severe fidelity loss at extreme perturbations (*σ* = 0.10, 24.29± 0.43 dB vs. 23.79 ± 0.08 dB). This enhanced resilience at high-variance regimes stems from our context-aware filtering and inductive morphological regularization.

**TABLE I:** Quantitative Benchmark on Held-Out Test Set (*N* = 9,908). Throughput Measured on a Single GPU With Batch Size *B* = 1.

| **Method** | **PSNR (dB)↑** | **SSIM↑** | **MSE↓** | **RMSE↓** | **Cosine Sim↑** | **LPIPS↓** | **FPS↑** |
|---|---|---|---|---|---|---|---|
| **MorphoStain (Ours)** | **40.56** | **0.9961** | **9.092 × 10<sup>−5</sup>** | **0.0095** | **0.9999** | 0.0141 | 140.1 |
| InverseAE (SOTA) [11] | 38.84 | 0.9934 | 1.349 × 10<sup>−4</sup> | 0.0115 | 0.9999 | 0.0150 | **197.2** |
| NMF [25] | 35.39 | 0.9927 | 2.904 × 10<sup>−4</sup> | 0.0170 | 0.9999 | **0.0140** | 16.9 |
| Ruifrok & Johnston [5] | 34.06 | 0.9896 | 4.079 × 10<sup>−4</sup> | 0.0200 | 0.9997 | 0.0499 | 18.8 |
| Macenko *et al.* [6] | 32.37 | 0.9880 | 5.821 × 10<sup>−4</sup> | 0.0241 | 0.9999 | 0.0165 | 17.4 |
| Sparse NMF [24] | 25.64 | 0.9448 | 2.736 × 10<sup>−3</sup> | 0.0523 | 0.9995 | 0.0743 | 16.1 |
| Vahadane *et al.* [7] | 21.40 | 0.8153 | 7.260 × 10<sup>−3</sup> | 0.0851 | 0.9987 | 0.1485 | 14.5 |
| LassoReg [27] | 18.37 | 0.5425 | 1.466 × 10<sup>−2</sup> | 0.1209 | 0.9971 | 0.2971 | 16.7 |

**TABLE II:** Ablation Study of Architectural Components and Distillation Dynamics Under Identical Single-GPU Calibration Conditions.

| **Configuration** | **Params (M)** | **Latency (ms)↓** | **FPS↑** | **PSNR (dB)↑** | **SSIM↑** | **RMSE↓** | **LPIPS↓** |
|---|---|---|---|---|---|---|---|
| M1: MorphoStain (Teacher) | 169.67 | 45.13 | 22.2 | 32.40 | 0.9903 | 0.0240 | 0.0483 |
| M2: No Phikon [16] (CNN Base) | 83.87 | 27.19 | 36.8 | 21.68 | 0.8462 | 0.0825 | 0.2649 |
| M3: No Physics Loss | 169.67 | 45.73 | 21.9 | 24.19 | 0.9301 | 0.0618 | 0.1660 |
| M4: No Overlap Penalty | 169.67 | 45.16 | 22.1 | 27.21 | 0.9701 | 0.0436 | 0.0964 |
| **MorphoStain (Student)** | **0.12** | **5.73** | **174.6** | **41.66** | **0.9965** | **0.0083** | **0.0318** |

**TABLE III:**
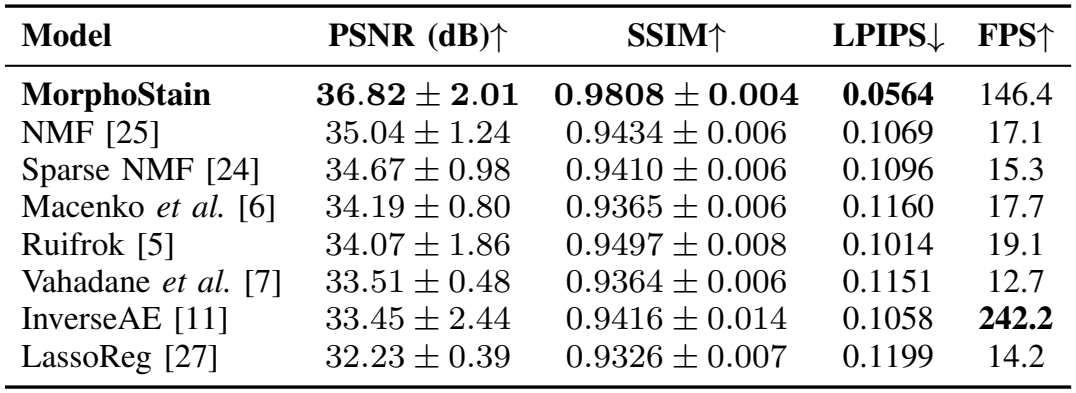
Zero-Shot Out-of-Domain Validation (*N* = 100) Evaluated Across 300 Deterministic Runs (3 Seeds) on Unseen Patches From Donor H14.09.010.

| **Model** | **PSNR (dB)↑** | **SSIM↑** | **LPIPS↓** | **FPS↑** |
|---|---|---|---|---|
| **MorphoStain** | **36.82 ± 2.01** | **0.9808 ± 0.004** | **0.0564** | 146.4 |
| NMF [25] | 35.04 ± 1.24 | 0.9434 ± 0.006 | 0.1069 | 17.1 |
| Sparse NMF [24] | 34.67 ± 0.98 | 0.9410 ± 0.006 | 0.1096 | 15.3 |
| Macenko *et al.* [6] | 34.19 ± 0.80 | 0.9365 ± 0.006 | 0.1160 | 17.7 |
| Ruifrok [5] | 34.07 ± 1.86 | 0.9497 ± 0.008 | 0.1014 | 19.1 |
| Vahadane *et al.* [7] | 33.51 ± 0.48 | 0.9364 ± 0.006 | 0.1151 | 12.7 |
| InverseAE [11] | 33.45 ± 2.44 | 0.9416 ± 0.014 | 0.1058 | **242.2** |
| LassoReg [27] | 32.23 ± 0.39 | 0.9326 ± 0.007 | 0.1199 | 14.2 |

**TABLE IV:** Robustness Under Illumination Drift Comparing Reconstruction Fidelity Between MorphoStain and InverseAE.

|  | **MorphoStain** |  |  | **InverseAE [11]** |  |
|---|---|---|---|---|---|
| **Shift Factor** | **PSNR↑** | **SSIM↑** | **MSE↓** | **PSNR↑** | **SSIM↑** |
| 1.00 (Nominal) | **40.75** | **0.9955** | 8.40 × 10<sup>−5</sup> | 39.30 | 0.9933 |
| 1.10 | **40.25** | **0.9946** | 9.40 × 10<sup>−5</sup> | 32.12 | 0.9863 |
| 1.20 | **38.22** | **0.9898** | 1.51 × 10<sup>−4</sup> | 26.81 | 0.9672 |
| 1.30 (Severe) | **35.37** | **0.9776** | 2.90 × 10<sup>−4</sup> | 23.73 | 0.9325 |

**Fig 4:**
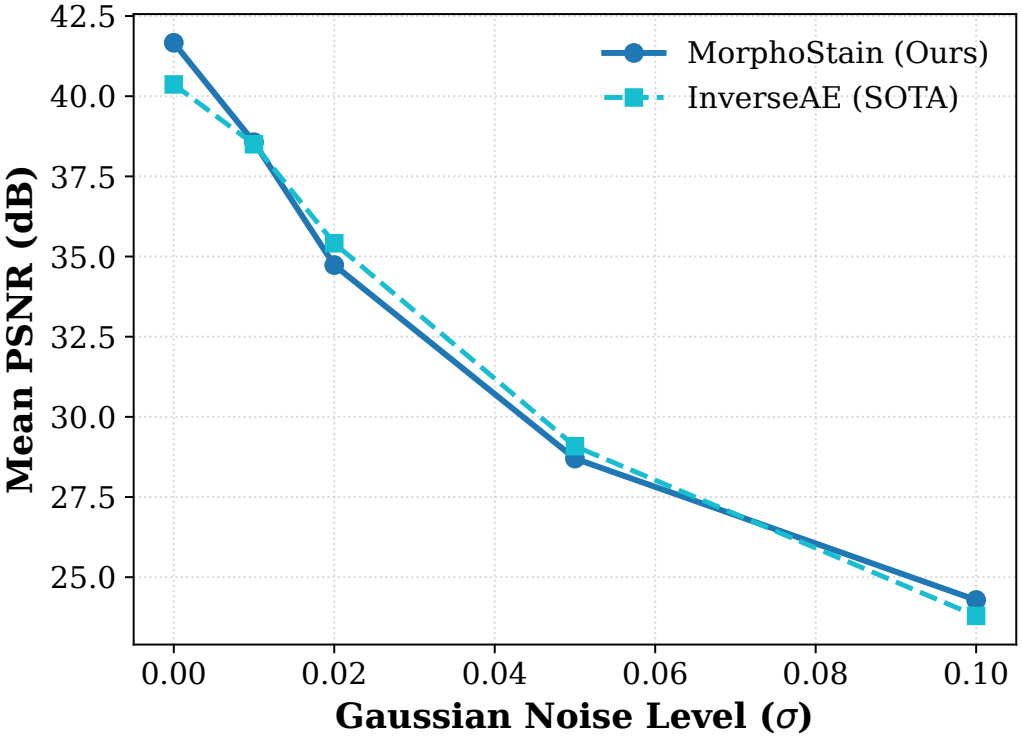
Degradation trajectories under additive Gaussian sensor noise. Mean PSNR across varying noise standard deviations (*σ* ∈ [0.0, 0.10]). MorphoStain maintains structural stability and prevents catastrophic numerical collapse under extreme optical corruption.

### E. Subgroup Analysis and Generalization

#### 1) Disease Burden and Subgroups

Partitioning by patho-logical burden demonstrates consistent reconstruction accuracy across spectra (41.79 ± 0.42 dB PSNR, 0.9969 SSIM in high-burden regimes vs. 41.15 ± 0.94 dB PSNR, 0.9954 SSIM in low-burden regimes; *N* = 200 each), alongside 41.47± 0.80 dB PSNR (0.9962 SSIM) across the *N* = 400 Tau (AT8) evaluation subset, reflecting robust numerical stability across biological density extremes. Furthermore, evaluation on a strictly balanced cohort (*N* = 100 per target) confirms uniform fidelity between Amyloid-*β* (40.80 ± 0.83 dB PSNR, 0.9972 ± 0.0004 SSIM) and Tau AT8 (40.12 ± 1.24 dB PSNR, 0.9946 ± 0.0016 SSIM) targets.

**TABLE V:**
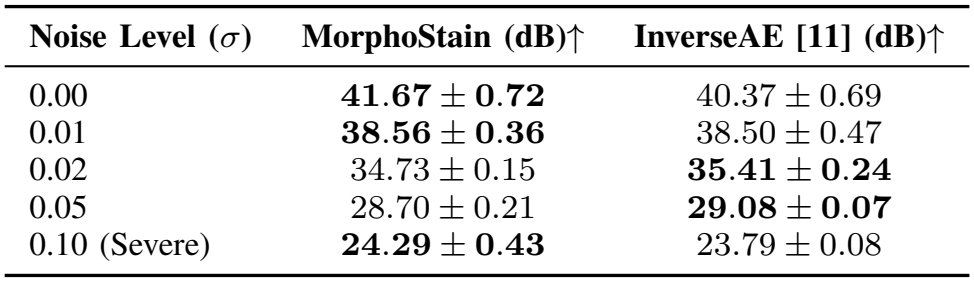
Additive Gaussian Sensor Noise Robustness (Re-construction PSNR Mean ± Std in dB).

#### 2) Cross-Tissue Transfer

In zero-shot deployments across human breast cancer IHC sections from the external BCI benchmark [33] (HER2/Hematoxylin) evaluated directly with-out color normalization, MorphoStain resolved cell membrane HER2 expression (39.12% ±16.13%) from dense HER2 (60.88% ±16.13%) and nuclear chromatin counterstain (30.07% 13.70%) while preserving tightly calibrated epistemic bounds (*u*_var_ = 4.68 ×10^−5^ ±2.19 ×10^−5^); see Supplementary Table S6 and Supplementary Fig. S7.

#### 3) Uncertainty Diagnostics and Failure Modes

Inspection of out-of-distribution reconstruction anomalies (PSNR *<* 35 dB under unscreened optical perturbations) reveals that fidelity drops correlate strictly with severe out-of-focus blurs and tissue folding boundaries. Crucially, the epistemic uncertainty head flags these artifactual zones (*u*_var_ *>* 0.0025), providing a reliable automated gating metric for clinical quality control (Fig. 5 and Supplementary Fig. S5).

**Fig 5:**
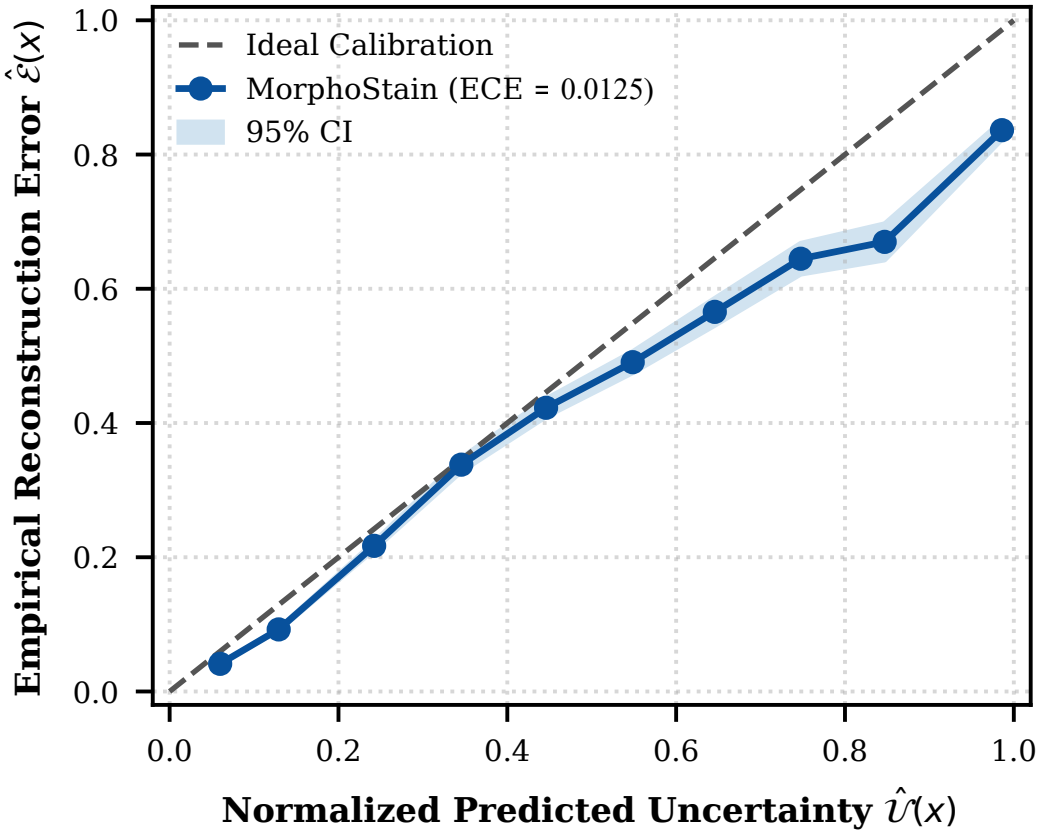
Reliability diagram and epistemic uncertainty calibration. Empirical reconstruction error (MSE) versus normalized predicted uncertainty *Û*(*x*) across test patches, achieving neardiagonal alignment (ECE = 0.0125).

## VI. Discussion, Limitations, and Future Work

### A. Physical Disentanglement and Distillation Dynamics

Classical color deconvolution methods [5], [6] suffer from severe crosstalk bleed when resolving spectrally identical chromogens, whereas unconstrained generative models [11] drift into ill-posed solutions without physical priors (Table II). MorphoStain bridges this gap by anchoring a Latent Diffusion Teacher within a Beer–Lambert OD manifold. By fixing the distilled student’s objectives to the analytical Moore– Penrose projection, it preserves exact spectral conservation and bypasses the high-frequency sampling variance inherent in iterative stochastic diffusion.

Furthermore, resolving dual-DAB mIHC in Alzheimer’s pathology [1] typically forces standard matrix factorization into a single uninformative channel. Our Symmetric Morpho-logical Routing Module (*M*_attn_) circumvents this by leveraging slide-level cohort metadata as a weakly supervised prior, enabling zero-shot, label-free biomarker separation. Finally, the epistemic uncertainty head (*u*_var_) provides an automated clinical safety gate; as shown in Fig. 5, predicted variance closely tracks local reconstruction error (ECE = 0.0125) to flag out-of-focus and boundary artifacts without human intervention.

### B. Limitations and Future Directions

Despite strong generalization across external cohorts, three operational limitations motivate future work:

1. **Static Calibration:** Extreme over-staining may violate nominal OD bounds. Future work will embed differentiable absorbance estimation within the Phikon ViT [16] for slide-specific stain adaptation.
2. **Active Triaging:** While *u*_var_ reliably flags optical defocus (Fig. 5), our pipeline relies on upstream tile rejection. We aim to integrate these maps directly into WSI scanners for real-time human-in-the-loop clinical triaging.
3. **Multiplexing Scope:** Currently targeting *M* = 2, extending *M*_attn_ to softmax-gated multi-head decoders for higher-order multiplex immunofluorescence (*M*≥ 4) remains an important direction.

## VII. Conclusion

MorphoStain introduces a physics-guided teacher–student framework that unifies Beer–Lambert constraints with pathology foundation models and latent diffusion to resolve the mathematically ill-posed stain decomposition problem. By distilling diffusion priors into a lightweight student anchored by morphological routing, the framework achieves SOTA realtime inference, separates overlapping identical chromogens without pixel-level annotations, and provides calibrated epistemic uncertainty for automated quality control. Extensive evaluations demonstrate significant statistical gains, zero-shot domain transferability, and robust resilience to optical shifts, establishing MorphoStain as a scalable and reliable tool for quantitative clinical neuropathology.

## Data Availability

The neuropathological whole-slide imaging data used in this study are publicly available through the SEA-AD Data Portal (https://portal.brain-map.org/explore/seattle-alzheimers-disease) and the AWS Open Data Registry (s3://sea-ad-quantitative-neuropathology/). The external breast cancer HER2 IHC dataset is publicly available via the BCI Challenge (http://bci.grand-challenge.org/). The source code, pre-trained weights, and interactive clinical demonstration suite for MorphoStain are publicly available at https://github.com/mrsaraei/MorphoStain.

https://portal.brain-map.org/explore/seattle-alzheimers-disease

http://bci.grand-challenge.org/](http://bci.grand-challenge.org/

## Ethics Statement

This study exclusively utilized publicly available, deidentified human neuropathological and breast cancer immunohistochemical whole-slide imaging datasets obtained from the Seattle Alzheimer’s Disease Brain Cell Atlas (SEA-AD) Data Portal and the Breast Cancer Immunohistochemical Image Generation (BCI) Challenge. All original tissue collections and data sharing protocols were approved by the respective institutional review boards (IRBs) of the originating consortia. Because no new human subjects were recruited and no identifiable patient information was accessed, this secondary computational analysis was exempt from additional IRB approval.

## Acknowledgment

The authors would like to thank the Seattle Alzheimer’s Disease Brain Cell Atlas (SEA-AD) consortium and the organizers of the Breast Cancer Immunohistochemical Image Generation (BCI) Challenge for making their datasets publicly available.

## Data and Code Availability

The neuropathological whole-slide imaging data used in this study are publicly available through the SEA-AD Data Portal (https://portal.brain-map.org/explore/seattle-alzheimers-disease) and the AWS Open Data Registry (s3://sea-ad-quantitative-neuropathology/) [1]. The external breast cancer HER2 IHC dataset is publicly available via the BCI Challenge (http://bci.grand-challenge.org/) [33]. The source code, pre-trained weights, and interactive clinical demonstration suite for MorphoStain are publicly available at https://github.com/mrsaraei/MorphoStain.

## Notes

Preprint version. This article has supplementary downloadable material provided by the authors.

### Competing Interest Statement

The authors have declared no competing interest.

