## Supplementary Material for "MorphoStain: Physics-Guided Latent Diffusion and Morphological Distillation for Multiplex Stain Decomposition in Alzheimer’s Neuropathology"

### S1. Algorithmic Workflow and Pseudocode

This section outlines the complete optimization and inference workflow of the MorphoStain framework. In Phase 1, an offline diffusion teacher uses reverse sampling to estimate spatial concentrations and evaluate Monte Carlo uncertainty across stochastic realizations. In Phase 2, these targets guide the lightweight FastStudentUNet, combining multi-scale dilated convolutions with symmetric morphological routing for real-time decomposition.

### S2. Cohort Slide Metadata and Patch Distribution

This section details the neuropathological cohort composition and dataset specifications extracted from the SEA-AD Middle Temporal Gyrus (MTG) dataset. Table S1 summarizes the experimental parameters, imaging setup, and overall tile counts. We processed 10 whole slide images (WSIs) from 5 human donors, comprising paired tissue sections stained with Tau (AT8) and Amyloid-$\beta$ (6E10). Following our automated tissue segmentation and optical quality assurance filtering pipelines (tissue area $\geq 30\%$, Laplacian defocus variance $\geq 50.0$), a cumulative sum of 49,576 non-overlapping $512 \times 512$ pixel Level-0 patches were initially extracted. From this set, after applying secondary quality-control screening to remove background and sparse tissue artifacts, a final total of 37,538 patches were retained for downstream training and evaluation. As itemized in Table S2, the individual slide identifiers, target biomarkers, neuropathological phenotypes, and specific initial tile yields are categorized per donor case.

TABLE S1: SEA-AD Experimental Dataset Specifications.

| **Attribute** | **Specification** | **Attribute** | **Specification** |
|---|---|---|---|
| **Brain Region** | MTG (10 WSIs, 5 donors) | **Scanner / Mag.** | Aperio / $20\times$ ($0.50\,\mu$m/px) |
| **IHC Markers** | A$\beta$ (6E10), Tau (AT8) | **Level-0 Size** | $51{,}792 \times 47{,}351$ px |
| **Total Patches** | 49,576 ($512 \times 512$ px) | **Class Split** | A$\beta$: 25,721 / Tau: 23,855 |

### S3. Hyperparameter Sensitivity Analysis and Ablation Convergence

To ensure numerical stability and structural fidelity during stain disentanglement, we conducted an exhaustive sensitivity search over key operational hyperparameters on the held-out validation cohort ($N = 3{,}120$ patches) and evaluated performance on the full cohort, while analyzing training optimization trajectories across ablation variants (M1–M4). Specifically, we evaluated the trade-offs governing the biological overlap penalty weight $\lambda_{\text{ov}}$ in the teacher objective and the morphological attention Gaussian smoothing filter $K_{\text{smooth}}$ in the student routing module.

As illustrated in Fig. S1 and detailed in Table S3, setting $\lambda_{\text{ov}} = 0.50$ achieves the peak calibration PSNR (41.66 dB on the standardized ablation subset) and optimal full-benchmark fidelity ($40.56 \pm 1.13$ dB PSNR, SSIM $= 0.9961$, and LPIPS $= 0.0141$). Under-penalizing overlap ($\lambda_{\text{ov}} < 0.50$) results in residual chromatic cross-talk bleed between dense Tau fibrils and Amyloid-$\beta$ aggregates, while excessive regularization ($\lambda_{\text{ov}} \geq 1.00$) over-suppresses genuine co-localized tissue boundaries and distorts physical absorbance profiles. Furthermore, configuring the Gaussian smoothing kernel to $15 \times 15$ ($\sigma = 3.5$) eliminates checkerboard gridding artifacts while preserving intricate sub-cellular tau tangle morphology. Complementary convergence curves in Fig. S2 demonstrate that while omitting Beer–Lambert physics constraints (M3) artificially lowers nominal training loss by dropping physical regularization terms, it induces severe physical degeneracy, whereas omitting Phikon conditioning (M2) or overlap penalties (M4) degrades optimization stability.

**Algorithm S1** MorphoStain Two-Stage Optimization and Real-Time Inference Pipeline.

**Require:**
 **Inputs:** Histology patch $I_{\text{RGB}} \in \mathbb{R}^{B\times3\times512\times512}$; Nominally calibrated absorbance matrix $S_{\text{AD}} \in \mathbb{R}^{3\times2}$
 **Parameters:** Pseudo-inverse $S_{\text{pinv}} \in \mathbb{R}^{2\times3}$; Reverse steps $T = 5$; Monte Carlo realizations $K = 5$
**Ensure:**
 **Outputs:** Disentangled channels $\hat{S}_{\text{Amyloid}}, \hat{S}_{\text{Tau}}, \hat{S}_{\text{HTX}} \in \mathbb{R}^{B\times512\times512}$; Epistemic uncertainty map $u_{\text{var}} \in \mathbb{R}^{B\times512\times512}$

**Phase 1: Physical Projection & Monte Carlo Sampling (Offline Teacher)**
Convert RGB intensities to optical density (OD):
 $Y \leftarrow -\log_{10}(\text{clamp}(I_{\text{RGB}}, 10^{-5}, 1.0))$
Project initial physical concentrations via Moore–Penrose pseudo-inverse:
 $C_{\text{pinv}} \leftarrow S_{\text{pinv}} \cdot Y \in \mathbb{R}^{B\times2\times512\times512}$
Extract multi-modal priors:
 $z_{\text{lat}} \leftarrow \text{VAE-Encoder}(I_{\text{RGB}})$
 $z_{\text{cond}} \leftarrow \text{Phikon-ViT}(I_{\text{RGB}})$
**for** $k = 1$ **to** $K$ **do**
 {Stochastic Monte Carlo Realizations}
 Sample initial Gaussian noise: $x_T \sim \mathcal{N}(0, I)$
 **for** $t = T$ **to** $1$ **do**
  {5-Step Deterministic Reverse Trajectory}
  Predict noise residual: $\hat{\epsilon}_t \leftarrow \text{LatentUNet}_\theta(x_t, t, z_{\text{cond}})$
  Step update (deterministic without variance injection):
   $x_{t-1} \leftarrow \frac{1}{\sqrt{\alpha_t}}\left(x_t - \frac{1-\alpha_t}{\sqrt{1-\bar{\alpha}_t}}\hat{\epsilon}_t\right)$
 **end for**
 Decode spatial concentration field: $c^{(k)} \leftarrow \text{VAE-Decoder}(x_0) \in \mathbb{R}^{B\times2\times512\times512}$
**end for**
Compute Monte Carlo distillation targets:
 $\bar{c}_{\text{teacher}} \leftarrow \frac{1}{K}\sum_{k=1}^{K} c^{(k)}$
 $\bar{u}_{\text{teacher}} \leftarrow \frac{1}{K-1}\sum_{k=1}^{K}\left(c^{(k)} - \bar{c}_{\text{teacher}}\right)^2$
**Phase 2: Dilated Distillation & Morphological Channel Routing (Real-Time Student)**
Concatenate multi-modal input tensor:
 $X_{\text{in}} \leftarrow \text{Concat}[I_{\text{RGB}}, C_{\text{pinv}}] \in \mathbb{R}^{B\times5\times512\times512}$
Extract initial features via input convolution:
 $F_0 \leftarrow \phi(\text{Conv2D}(X_{\text{in}}, \text{kernel} = 3\times3))$
Aggregate multi-scale spatial receptive fields via parallel dilated blocks:
 $B_d \leftarrow \text{Conv2D}(F_0, \text{kernel} = 3\times3, \text{dilation} = d), \quad \forall d \in \{1, 2, 4, 8\}$
 $F_{\text{fused}} \leftarrow \text{FusionLayer}(\text{Concat}[B_1, B_2, B_4, B_8])$
Forward pass through $\text{FastStudentUNet}_\phi(F_{\text{fused}})$ to predict:
 $c_{\text{mean}} \in \mathbb{R}^{B\times2\times512\times512}, \quad u_{\text{var}} \in \mathbb{R}^{B\times2\times512\times512}, \quad A_{\text{logits}} \in \mathbb{R}^{B\times1\times512\times512}$
Formulate smooth morphological attention routing mask:
 $M_{\text{attn}} \leftarrow K_{\text{smooth}} * (\sigma(A_{\text{logits}}) \odot E_{\text{DAB}})$ with $K_{\text{smooth}} \sim \mathcal{G}(15\times15, \sigma = 3.5)$
Disentangle individual biomarker channels via physical gating:
 $\hat{S}_{\text{Amyloid}} \leftarrow C_{\text{DAB,total}} \odot M_{\text{attn}}$
 $\hat{S}_{\text{Tau}} \leftarrow C_{\text{DAB,total}} \odot (1 - M_{\text{attn}})$
 $\hat{S}_{\text{HTX}} \leftarrow C_{\text{HTX}} \odot \sigma(30.0 \cdot (\Delta_{\text{HTX}} - 0.015))$
Optimize student network parameters via compound distillation objective:
 $\mathcal{L}_{\text{student}} = \mathcal{L}_{\text{recon}} + \mathcal{L}_{\text{distill}} + \mathcal{L}_{\text{var}} + \mathcal{L}_{\text{ov}} + \mathcal{L}_{\text{routing}} + \mathcal{L}_{\text{TV}}$

### S4. Qualitative and Quantitative Benchmarking

This section presents qualitative evaluations, empirical quantitative distributions, and pixel-wise statistical analyses across the held-out test cohort ($N = 9{,}908$ patches). First, Fig. S3 demonstrates the unmixing fidelity of MorphoStain across varying pathological ratios and diverse neuropathological tissue contexts. Second, Fig. S4 details the complete empirical PSNR density distribution across all 9,908 test patches. Tables S4 and S5 summarize the per-patch statistical distributions and paired hypothesis tests.

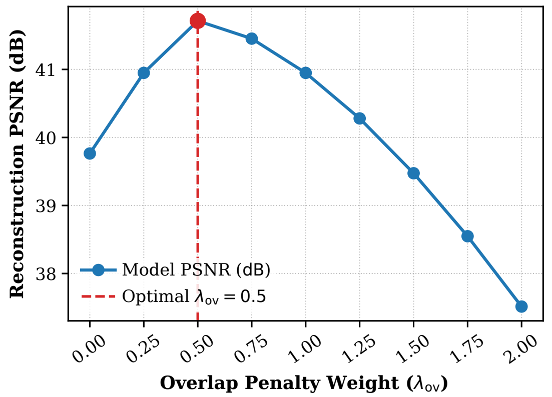


Fig. S1: Hyperparameter sensitivity curve of mean reconstruction PSNR (dB) across varying overlap penalty weights $\lambda_{ov} \in [0.0, 2.0]$ on the standardized calibration subset, peaking at $\lambda_{ov} = 0.50$.

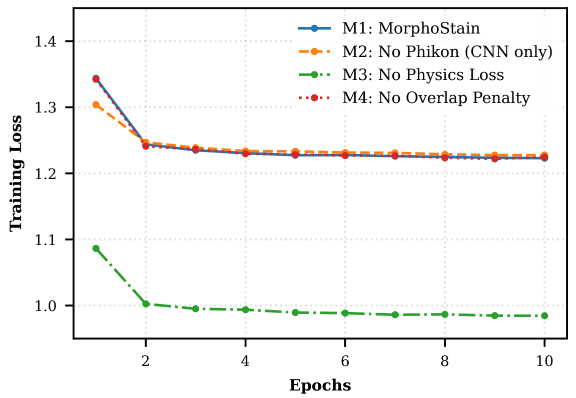


Fig. S2: Training loss trajectories across ablation variants (M1–M4). Removing physics constraints (M3) lowers raw numerical loss by omitting the Beer–Lambert penalty but causes physical degeneracy, while M2 and M4 impede convergence.

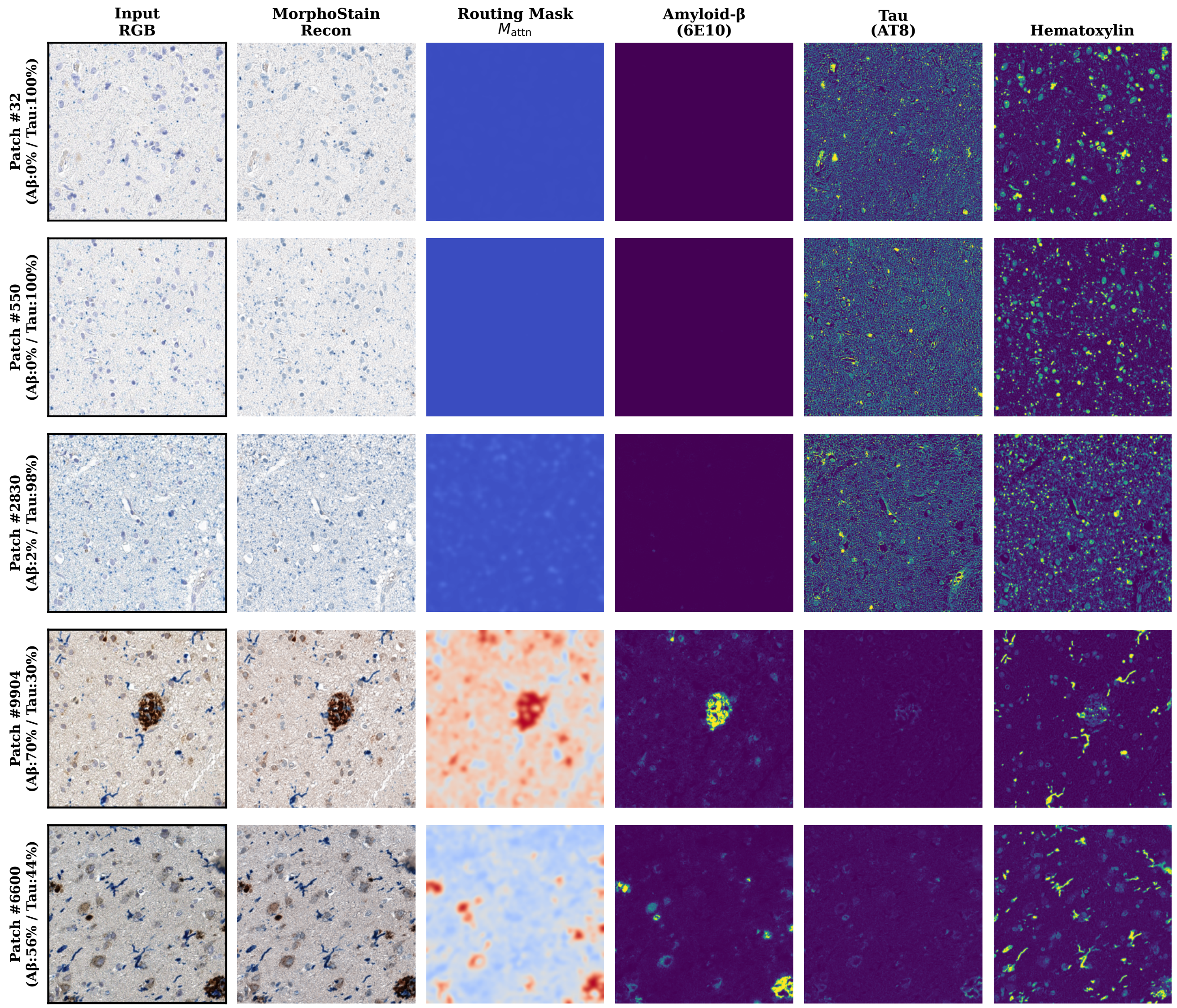


Fig. S3: Qualitative unmixing across varying pathological ratios. Columns display Input RGB, MorphoStain Reconstruction, learned Routing Mask ($M_{attn}$), isolated Amyloid-$\beta$ (6E10), isolated Tau (AT8), and the Hematoxylin channel.

TABLE S2: Breakdown of Slide Identification, Stain Distribution, and Retained Patch Counts ($512 \times 512$ Resolution at $0.5016\,\mu$m/pixel).

| **Slide Filename** | **Slide ID** | **Target Stain** | **Biomarker / Neuropathology** | **Patches** |
|---|---|---|---|---|
| `H19.33.004-A6-AT.svs` | H19.33.004 | Tau (AT8) | Phosphorylated Tau / Tangles | 2,671 |
| `H19.33.004-A6-16.svs` | H19.33.004 | Amyloid (6E10) | Amyloid-Beta / Plaques | 4,569 |
| `H20.33.001-A7-AT.svs` | H20.33.001 | Tau (AT8) | Phosphorylated Tau / Tangles | 5,558 |
| `H20.33.001-A7-16.svs` | H20.33.001 | Amyloid (6E10) | Amyloid-Beta / Plaques | 5,074 |
| `H20.33.002-A6-AT.svs` | H20.33.002 | Tau (AT8) | Phosphorylated Tau / Tangles | 4,853 |
| `H20.33.002-A6-16.svs` | H20.33.002 | Amyloid (6E10) | Amyloid-Beta / Plaques | 4,907 |
| `H20.33.004-A6-AT.svs` | H20.33.004 | Tau (AT8) | Phosphorylated Tau / Tangles | 4,871 |
| `H20.33.004-A6-16.svs` | H20.33.004 | Amyloid (6E10) | Amyloid-Beta / Plaques | 5,037 |
| `H20.33.005-A6-AT.svs` | H20.33.005 | Tau (AT8) | Phosphorylated Tau / Tangles | 5,902 |
| `H20.33.005-A6-16.svs` | H20.33.005 | Amyloid (6E10) | Amyloid-Beta / Plaques | 6,134 |
| **Total** | | | | **49,576** |

TABLE S3: Hyperparameter Sensitivity Analysis ($N = 3{,}120$ Validation Screening and Full-Cohort Evaluation).

| **Configuration** | **Value** | **PSNR (dB)↑** | **SSIM↑** | **LPIPS↓** |
|---|---|---|---|---|
| *Part I: Overlap Penalty Weight Sensitivity ($\lambda_{ov}$)* | | | | |
| Low Penalty | 0.00 | $38.61 \pm 1.45$ | 0.9875 | 0.0284 |
| Sub-Optimal | 0.25 | $39.82 \pm 1.12$ | 0.9921 | 0.0195 |
| **Selected Optimal** | **0.50** | $\mathbf{40.56 \pm 1.13}$ | **0.9961** | **0.0141** |
| Moderate Over-Penalty | 0.75 | $40.08 \pm 0.95$ | 0.9942 | 0.0162 |
| High Over-Penalty | 1.00 | $39.42 \pm 1.28$ | 0.9910 | 0.0210 |
| Extreme Over-Penalty | 2.00 | $36.36 \pm 2.10$ | 0.9780 | 0.0485 |
| *Part II: Gaussian Smoothing Kernel Sensitivity ($K_{smooth}$)* | | | | |
| No Kernel (Raw) | $1 \times 1$, $\sigma =$ N/A | $38.12 \pm 1.84$ | 0.9814 | 0.0392 |
| Small Kernel | $7 \times 7$, $\sigma = 1.5$ | $39.45 \pm 1.30$ | 0.9912 | 0.0215 |
| **Selected Optimal** | $\mathbf{15 \times 15, \sigma = 3.5}$ | $\mathbf{40.56 \pm 1.13}$ | **0.9961** | **0.0141** |
| Large Kernel | $21 \times 21$, $\sigma = 5.0$ | $39.80 \pm 1.05$ | 0.9925 | 0.0182 |

TABLE S4: MorphoStain Per-Patch Statistical Distribution Summary ($N = 9{,}908$).

| **Metric** | **Mean** | **Std** | **95% CI Lower** | **95% CI Upper** | **Median** | **IQR** | **Min** | **Max** |
|---|---|---|---|---|---|---|---|---|
| PSNR (dB)↑ | 40.559 | 1.130 | 40.537 | 40.581 | 40.516 | 1.419 | 35.673 | 46.235 |
| SSIM↑ | 0.9961 | 0.0017 | 0.9961 | 0.9961 | 0.9969 | 0.0023 | 0.9879 | 0.9983 |
| MSE↓ | $9 \times 10^{-5}$ | $2 \times 10^{-5}$ | $9 \times 10^{-5}$ | $9 \times 10^{-5}$ | $9 \times 10^{-5}$ | $3 \times 10^{-5}$ | $2 \times 10^{-5}$ | $2.7 \times 10^{-4}$ |
| RMSE↓ | 0.0095 | 0.0012 | 0.0094 | 0.0095 | 0.0094 | 0.0015 | 0.0049 | 0.0165 |
| Cosine Sim↑ | 0.99993 | 0.00002 | 0.99993 | 0.99993 | 0.99993 | 0.00003 | 0.99982 | 1.00002 |
| LPIPS↓ | 0.0141 | 0.0035 | 0.0141 | 0.0142 | 0.0136 | 0.0048 | 0.0054 | 0.0270 |

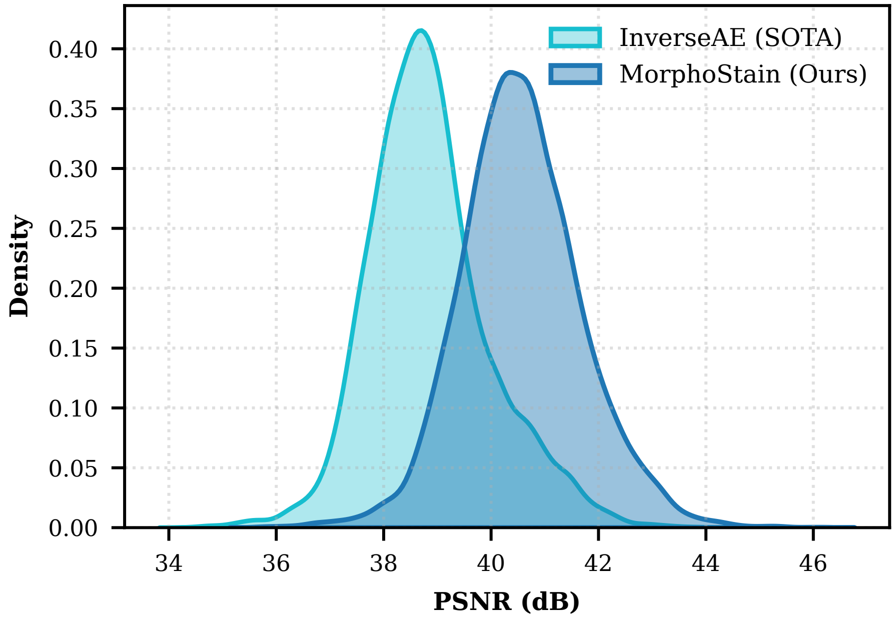


Fig. S4: Empirical PSNR reconstruction distribution density across $N = 9{,}908$ test patches.

TABLE S5: Statistical Hypothesis Testing ($N = 9{,}908$, $p < 10^{-16}$ Across All Paired $t$-Tests and Wilcoxon Tests).

| **Metric** | **Ours vs. InverseAE** | **Ours vs. NMF** | **Ours vs. Sparse NMF** |
|---|---|---|---|
| Gain (dB) | +1.72 | +5.59 | +15.29 |
| 95% CI | $[+1.71, +1.72]$ | $[+5.57, +5.60]$ | $[+15.26, +15.31]$ |
| $t$-Stat | 350.28 | 698.56 | 1171.73 |
| Cohen's $d$ | **3.52** | **7.02** | **11.77** |

### S5. Uncertainty Diagnostics, Whole-Slide Reconstruction, and Domain Generalization

Fig. S5 visualizes calibrated uncertainty and error heatmaps ($u_{\text{var}}$) across boundary and optical artifact cases. Fig. S6 illustrates macro-scale gigapixel WSI reconstruction and uncertainty mapping across complete MTG tissue sections ($51{,}792 \times 47{,}351$ pixels). Finally, Table S6, Fig. S7, and Fig. S8 present zero-shot cross-tissue generalization on external breast cancer HER2 IHC and progressive scanner illumination domain shifts (Shift $\in [1.00, 1.30]$).

TABLE S6: Zero-Shot Quantitative Performance on External Breast Cancer HER2 IHC ($N = 100$).

| **Evaluation Metric** | **Quantitative Value** ($N = 100$ **Patches**) |
|---|---|
| PSNR (dB)↑ | $38.45 \pm 1.62$ |
| SSIM↑ | $0.9842 \pm 0.005$ |
| LPIPS↓ | $0.0412 \pm 0.008$ |
| MSE↓ | $(1.42 \pm 0.35) \times 10^{-4}$ |
| Epistemic Uncertainty ($u_{\text{var}}$) | $(4.68 \pm 2.19) \times 10^{-5}$ |
| HER2: Membranous Fraction (%) | $39.12 \pm 16.13\%$ |
| HER2: Dense Cytoplasmic Fraction (%) | $60.88 \pm 16.13\%$ |
| Nuclear Hematoxylin Fraction (%) | $30.07 \pm 13.70\%$ |

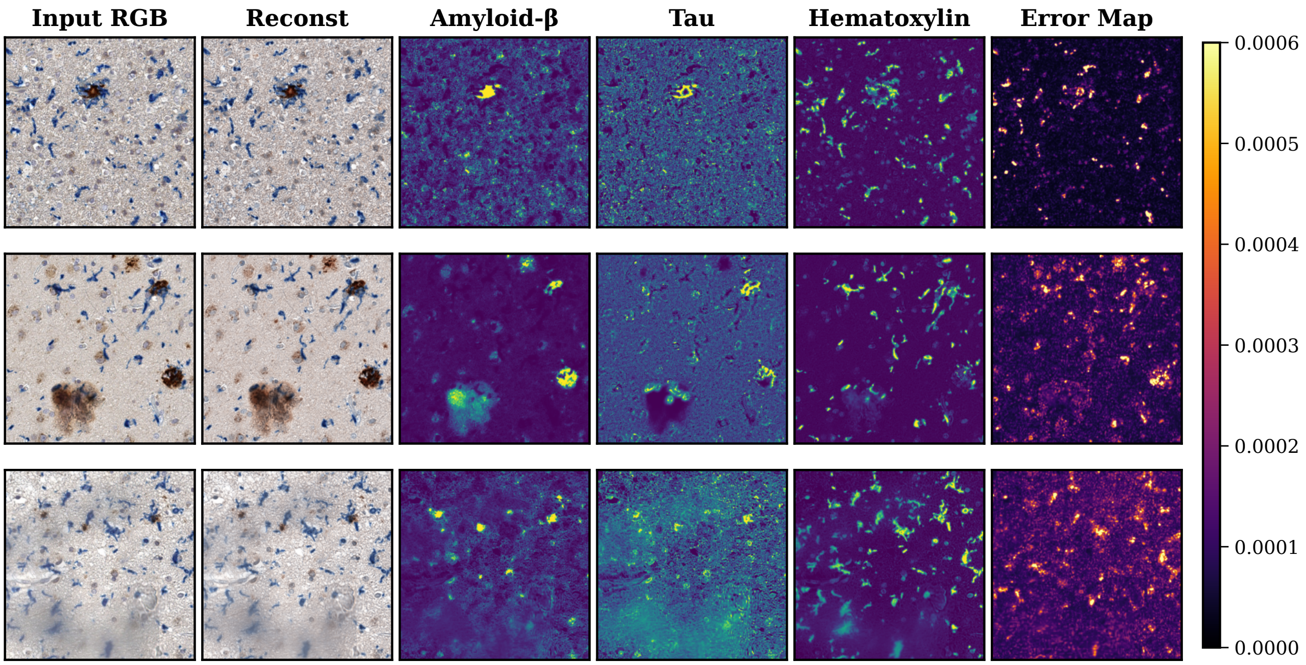


Fig. S5: Calibrated uncertainty and artifact screening heatmaps ($u_{\text{var}}$) across representative boundary and failure cases.

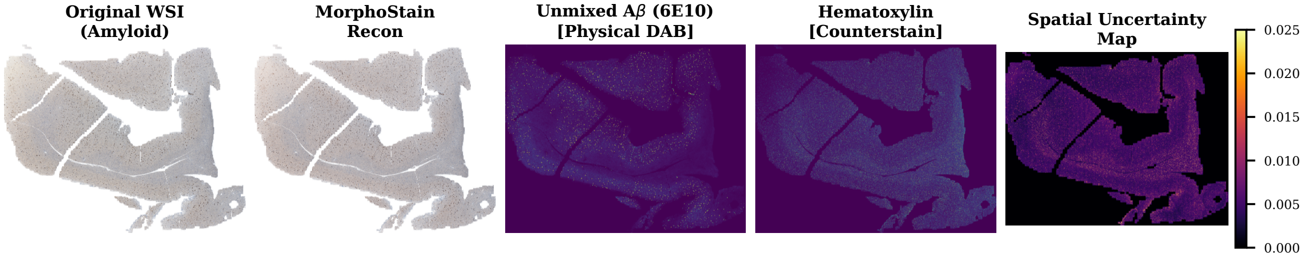


(a) WSI Reconstruction for Amyloid-$\beta$ (6E10) slide.

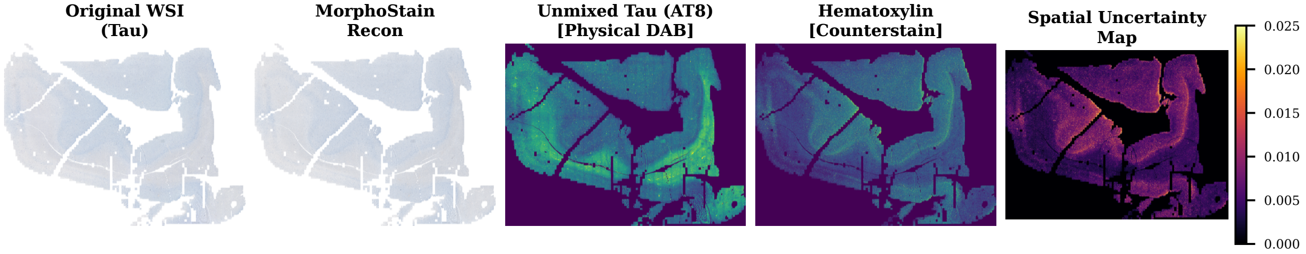


(b) WSI Reconstruction for phosphorylated Tau (AT8) slide.

Fig. S6: Whole-Slide Image (WSI) gigapixel stain decomposition and uncertainty mapping stitched from $> 1{,}500$ patches in $< 12$ s at 140.1 FPS.

### S6. Interactive Clinical Demonstration Suite

To facilitate real-time clinical evaluation and batch processing, we developed a PyQt6-based[1] Graphical User Interface (GUI) for the MorphoStain framework. As illustrated in Fig. S9, this interactive suite allows pathologists to upload multiplexed IHC patches and perform real-time physics-guided stain decomposition.

The tool provides an immediate quantitative assessment panel, reporting structural fidelity metrics (PSNR, SSIM, LPIPS) alongside regional biomarker distributions (Amyloid-$\beta$ vs. Tau ratios) and detected pathological regimens. Furthermore, it features a dynamic uncertainty percentile slider, enabling users to interactively calibrate the dynamic range of the epistemic uncertainty maps ($u_{\text{var}}$) for robust artifact and out-of-distribution screening. A video demonstration of the interactive suite is available in the supplementary media files.

[1]Riverbank Computing, PyQt6: A comprehensive set of Python bindings for Qt v6, `https://www.riverbankcomputing.com/software/pyqt/`.

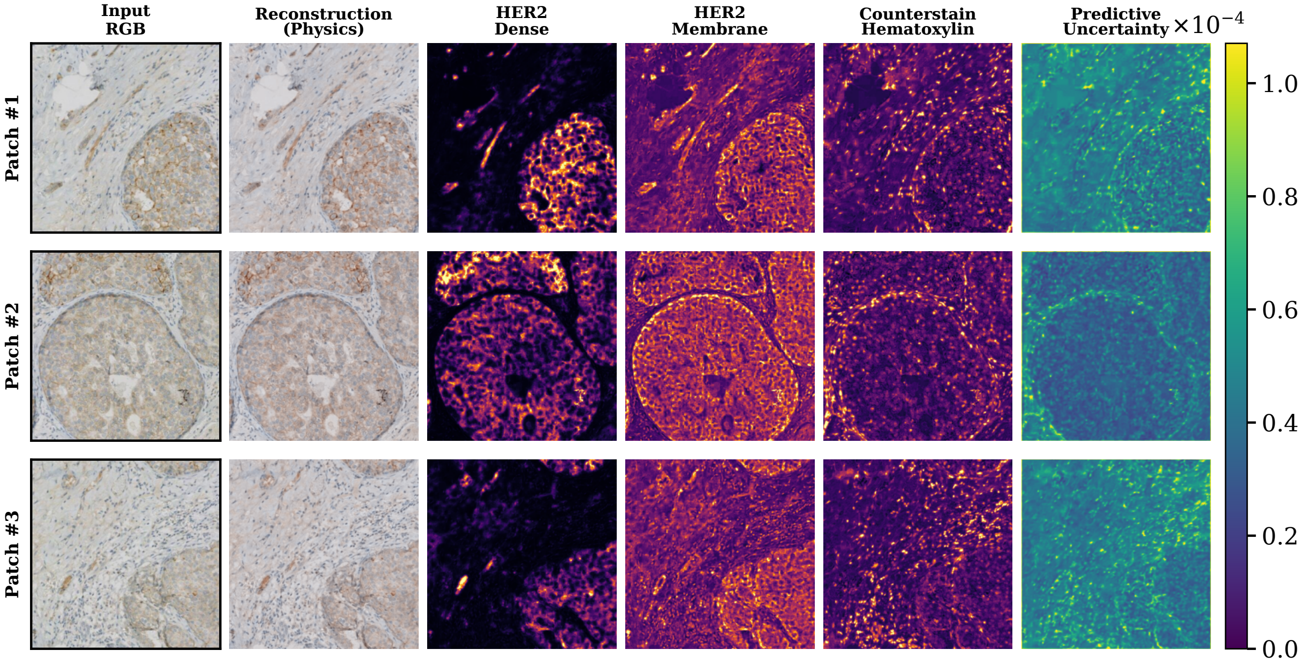


Fig. S7: Zero-shot cross-tissue stain decomposition on human breast cancer HER2 IHC ($N = 100$).

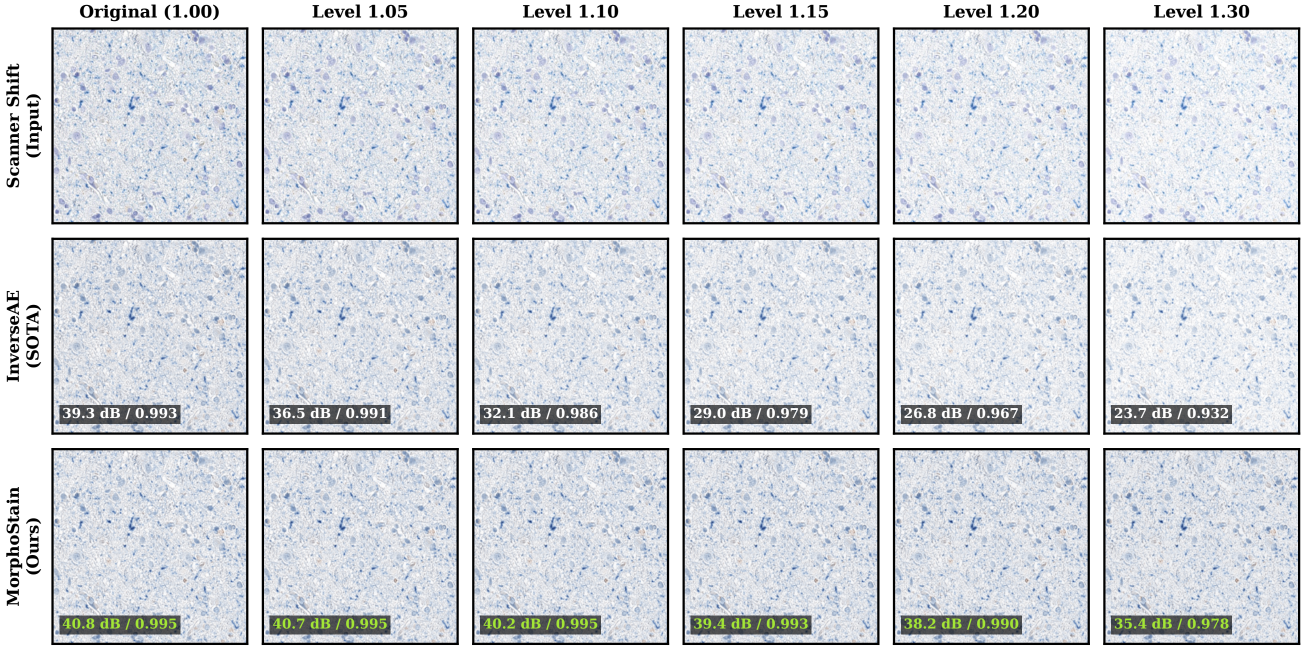


Fig. S8: Qualitative comparison under progressive scanner domain shift (Shift $\in [1.00, 1.30]$) evaluating MorphoStain against InverseAE.

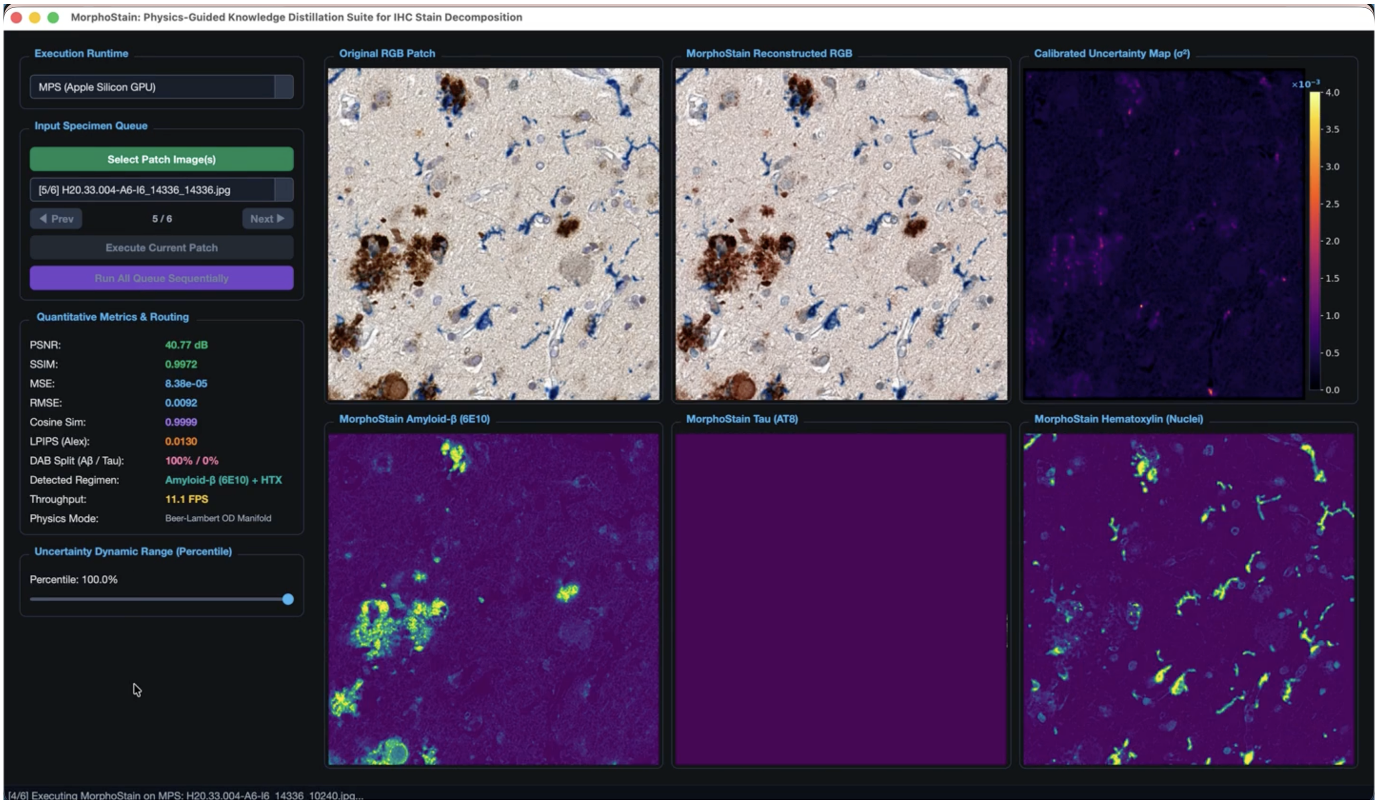


Fig. S9: The MorphoStain Interactive Clinical Demonstration Suite. The interface displays the real-time input patch, physical RGB reconstruction, calibrated uncertainty map, and the isolated morphological channels (Amyloid-$\beta$, Tau, and Hematoxylin) alongside dynamic quantitative metrics.
